# SARS-CoV-2 in wastewater treatment plants

**DOI:** 10.1101/2020.06.04.20122218

**Authors:** Maria Cristina Collivignarelli, Carlo Collivignarelli, Marco Carnevale Miino, Alessandro Abbà, Roberta Pedrazzani, Giorgio Bertanza

**Affiliations:** Department of Civil Engineering and Architecture, University of Pavia, via Ferrata 1, 27100 Pavia, Italy; Interdepartmental Centre for Water Research, University of Pavia, Via Ferrata 3, 27100 Pavia, Italy; Department of Civil, Environmental, Architectural Engineering and Mathematics, University of Brescia, Via Branze 43, 25123 Brescia, Italy; Department of Mechanical and Industrial Engineering, University of Brescia, via Branze 38, I-25123 Brescia, Italy

**Keywords:** Coronavirus removal, CoViD, Human health, SARS, Sewage sludge, Wastewater

## Abstract

As for the SARS coronavirus in the 2003 epidemic, the presence of SARS-CoV-2 has been demonstrated in faeces and, in some cases, urine of infected people, as well as in wastewater. This paper proposes a critical review of the state of the art regarding studies on the presence of SARS-CoV-2 in wastewater and sewage sludge, the factors affecting its inactivation and the main proposed treatments, with the aim to provide useful information at operative level in order to better and safer manage wastewater and sewage sludge. Given the lack of literature on SARS-CoV-2, studies involving other HCoVs such as SARS-CoV and HCoV-229E have been also considered. In wastewater, the resistance of SARS-CoV has proven to be very limited, especially at temperatures above 20 °C, and the virus has been easily removed with the use of chlorine (> 0.5 mg L^−1^ for 30 min). For sewage sludge, based on in vitro experiments, it is suggested to increase the retention times before a possible reuse in agriculture only for precautionary purposes, since SARS-CoV-2 is unlikely to occur in the sludge. SARS-CoV-2 in wastewater might track the epidemic trends: although being extremely promising, an effective and wide application of this approach requires a deeper knowledge of the amounts of viruses excreted through the faeces and the actual detectability of viral RNA in sewage.

## 1. Introduction

The coronavirus disease 2019 (CoViD-19) is caused by the virus capable of causing Severe Acute Respiratory Syndrome Coronavirus 2 (SARS-CoV-2), previously provisionally named 2019 novel coronavirus or 2019-nCoV (Gorbalenya, Alexander et al., 2020; Lai et al., 2020; Pal et al., 2020; X.-W. Xu et al., 2020). SARS-CoV-2 was discovered in December 2019 in Wuhan (Hubei, China) (Huang et al., 2020; Zhu et al., 2020) and quickly spread worldwide thanks to the high rate of infectivity (Remuzzi and Remuzzi, 2020) and the presence of a huge number of asymptomatic or minimally symptomatic patients, hardly identifiable (Zou et al., 2020). In March 2020, the World Health Organization declared a pandemic state (WHO, 2020a) and, despite in many countries total lockdown was imposed (Collivignarelli et al., 2020; Lau et al., 2020), CoViD-19 spread in more than 240 countries (25^th^ May, 2020), millions of people got infected and hundreds of thousands died (WHO, 2020b).

The transmission of SARS-CoV-2 person-to-person has been found to be due to its presence in the respiratory tract of infected people (Ghinai et al., 2020). SARS-CoV-2, however, has been also found in the faeces and urine of infected people, thus raising the questions about a possible oral/faecal transmission. This finding represents the starting point of two main fields of investigations: 1) the need of detecting and quantifying the number of viral particles and the actual pathogenicity in wastewater (WW) and sewage sludge (SS) produced by hospitals and houses with infected people, being a possible source of contamination (Núñez-Delgado, 2020); 2) the need of identifying the best strategies for treating/managing wastewater and residues; 3) the possible application of the waste based epidemiology (WBE) approach, in order to track the spatial and temporal dynamics of the epidemic and to get an early warning in case of future outbreaks (Daughton, 2020; Sims and Kasprzyk-Hordern, 2020). Since SARS-CoV-2 appeared recently, research in this field is in its infancy and the position of the scientific community is still uncertain. However, some studies on the behaviour of SARS-CoV and other coronaviruses conducted in recent years (Casanova et al., 2009; Gundy et al., 2009; Wang et al., 2005), may help to understand and predict the potential problems due by the presence of SARS-CoV-2 in the faeces and urine of infected subjects.

This paper reviews the available data about the presence of SARS-CoV-2 (together with other viruses, as indicators) in faces and urine, wastewater and sewage sludge, factors promoting its inactivation and possible treatments.

## 2. Bibliometric research and literature trends

At first, the presence of SARS-CoV-2 in faeces and urine was studied based on the very recent literature in this field. Since there are still few results published in the literature to ascertain the presence of SARS-CoV-2 in WW and there are no results on SS, the knowledge acquired so far for other types of coronavirus has been reviewed and discussed.

In order to carry out the review according to the objectives described above, a multi-step methodology has been adopted as reported by other authors (Collivignarelli et al., 2019; Kable et al., 2012; Martínez Fernández et al., 2019). Scopus® database has been used to search relevant literature research papers, reviews, books and conference proceedings. The keywords are “coronavirus”, “coronavirus AND faeces”, “coronavirus AND urine”, and “coronavirus AND wastewater”. The analysis has been carried out searching the keywords in the fields “Article title, Abstract, Keywords”. These papers have been checked in order to eliminate duplicates and out-of-scope documents. These data were also used to provide a chronological and spatial bibliometric analysis.

In recent years, while the number of publications related to coronaviruses reached significant levels thanks to the increased interest following the SARS and, in particular, CoViD-19 epidemic (more than 4900 in the first 4 months of 2020 only), the number of published studies on the presence and resistance of coronaviruses in WW is currently almost limited (only 13 from 2000 till now) (Fig. 1). This shortage becomes even more evident if studies on the presence of coronavirus in SS are quantified. Using “coronavirus AND sludge” in Scopus®, only 4 results were identified. The limited amount of studies contrasts with the collection of publications relating to the presence of Coronavirus in faeces and urine, for which greater interest has been shown (in the last 5 years there have been 353 and 52, respectively).

The USA and China have a prominent position in the list, followed by the United Kingdom and Germany. Italy, Canada, Japan, Hong Kong and Saudi Arabia, which appear among the most prolific nations in terms of publications in this field. Regarding the distribution of papers in the world, in case of co-authoring from institutions of different Countries, a ranking has been drawn up, considering the origin of the first author. The research paper category is always the most represented, with peaks of 76% –90% for publications about the presence of SARS-CoV-2 in faeces and urine.

**Fig. 1:**
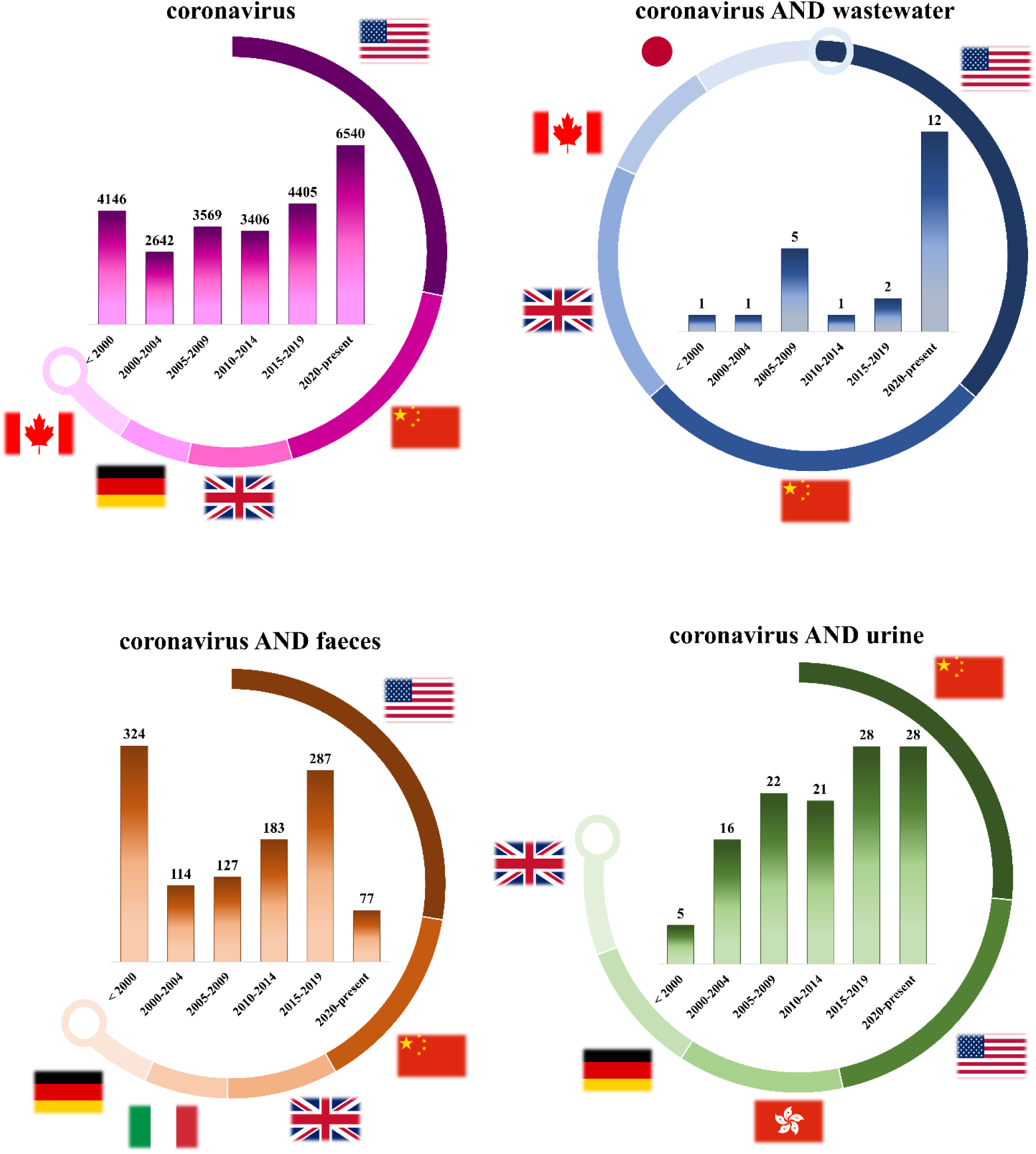
*Comparison of the trends of research papers, reviews, books and conference proceedings published up to 20^th^ May 2020 and their distribution in the world. All data were obtained introducing the words “coronavirus”, “coronavirus AND wastewater”, “coronavirus AND faeces”, and “coronavirus AND urine” in Scopus*^®^ *[two-column fitting image]*

## 3. Presence of SARS-CoV-2 in faeces and urine

Several studies have shown the presence of the SARS-CoV-2 in faeces and urine of infected patients (Chen et al., 2020; Gu et al., 2020; Huang et al., 2020; Tian et al., 2020; Y. Wu et al., 2020; Xiao et al., 2020). In Table 1, the results of recent clinical studies are reported. There is literature agreement about the possible finding in faeces of infected subjects, while the occurrence in the urine has not been always confirmed. For instance, Lescure et al. (2020) followed five patients admitted to French hospitals. In this case, the virus was detected in the faeces of two patients while no urine samples appeared positive.

From a critical analysis of literature, some fundamental aspects can be highlighted. The first concerns the low level of SARS-CoV-2 RNA shed in the first phase, after illness onset, demonstrated by a rRT-PCR (Real-Time Reverse Transcriptase-Polymerase Chain Reaction) cycle (C_t_) value generally in a range between 30 and 40 (considering 40 as positivity threshold) (Wang et al., 2020; Zhang et al., 2020), quite higher than that deriving from swabs of the respiratory tract (> 20) (Zou et al., 2020). While the viral load in the respiratory tract tends to run out more quickly, several studies have demonstrated that the virus can be found in the stool of an infected subject even after a prolonged period (Cai et al., 2020; Lescure et al., 2020). Zhang et al. (2020) observed that the median duration of virus emission was 10 days in swabs from respiratory tracts and up to 22 days in faeces. Despite the negativity of respiratory swabs, in 5% of cases, the faeces were tested positive also 26 days after discharge. Gupta et al. (2020) examined 26 articles on the presence of SARS-CoV-2 in faeces and revealed its occurrence up to 30 days after the onset of the first symptoms in infected patients (in a low number of cases, up to 47 days).

Although, at the moment, a path of oral-faecal contamination has not been demonstrated yet and further studies are required, this aspect is still very important because it could determine a greater probability of the presence of viral fragments in WW, also in areas where the infection is still in its early stages, thus allowing to follow the approach of the wastewater based epidemiology.

**Table 1:**
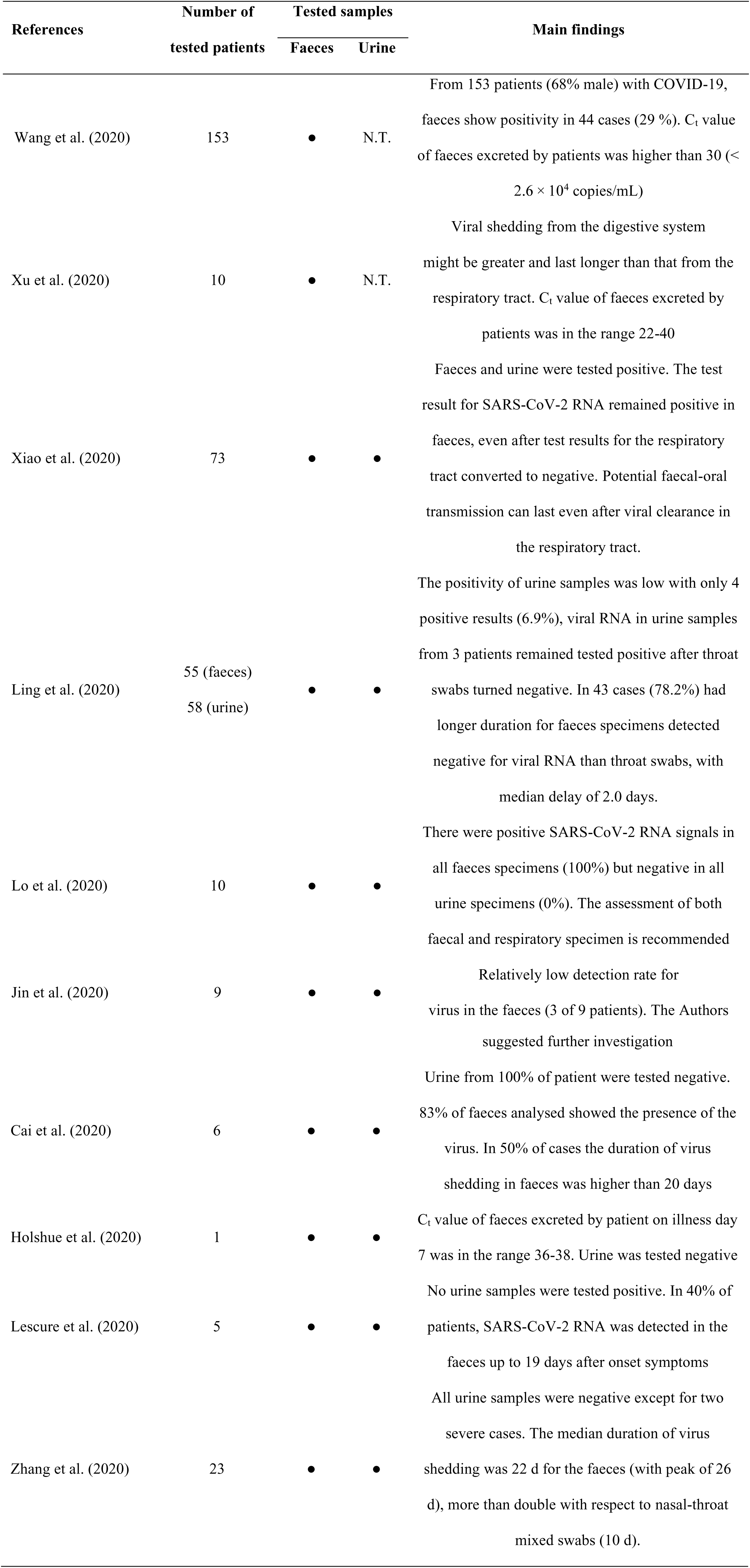
*Literature results on presence of SARS-CoV-2 in faeces and urine. N.T.: not tested [two-column fitting table]*

### 4.1. SARS-CoV 2 and wastewater

#### 4.1.1. Is the presence of the virus in wastewater possible?

The answer entails the availability of a standard technique for extract and quantify the virus in such a complex matrix, and, in particular, to assess its pathogenicity. As reported above, SARS-CoV-2 can reach the sewerage via faeces, and, possibly, urine, beside other human excreta (Table 2). However, the survival capacity of the virus in WW remains very uncertain and, to date, the results available on the SARS-CoV-2 are limited. Ahmed et al. (2020) researched the presence of the virus in the WW of the city of Brisbane (Australia) in the period at the time of the epidemic peak. Although it was detected in two samples, the quantification cycle (C_q_) values were very high (37.5 and 39, respectively) and comparable to those that the virus has in the faeces and urine. F. Wu et al. (2020) monitored a WWTP in Massachusetts determining the presence of SARS-CoV-2 RNA in an order of magnitude estimate of approximately 100 viral particles mL^−1^ of WW. In the Netherlands, Medema et al. (2020) studied the WW entering 8 different wastewater treatment plants (WWTPs), finding viral fragments of SARS-CoV-2 RNA. During the epidemic peak, Wurtzer et al. (2020) measured to 10^6^ – 10^7^ unit_SARS-CoV-2 RNA_ mL^−1^ in French sewage, while Randazzo et al. (2020b, 2020a) monitored the viral RNA in WW collected from two Spanish regions. The latter, quantified SARS-CoV-2 (5.38 ± 0.21 log genomic copies L^−1^) also in samples collected in La Murgia, an area, where CoViD-19 had a very low impact. Between February and April, in Milan and Rome 12 samples were collected from WWTPs and analysed by La Rosa et al. (2020b). SARS-CoV-2 RNA was evidenced in 50% of samples. Two aspects are particularly noteworthy in study: first, viral fragments were detected in Milan only few days after the first Italian case, was officially released (in the town of Codogno, about 60 km South of Milan). As suggested by La Rosa et al. (2020b) this means that the analysis of WW may lead to a more accurate estimation of the actual period of CoViD-19 onset. Secondly, they started from the standard WHO procedure for Poliovirus surveillance (WHO, 2003) and designed a new SARS-CoV-2 specific primer set for molecular analysis, in order to search the SARS-CoV-2. The development of a standardized procedure of SARS-CoV-2 RNA detection in WW is a crucial aspect in order to obtain reliable and comparable results. However, to date there is no a univocal method, and this represents a significant gap in the research, regarding both the phases of extraction (from such a complex matrix) and quantification.

These studies clearly confirm that the virus can therefore be present in WW, but many points remain uncertain. For instance, it is mandatory to investigate the impact of external conditions, such as temperature, pH and retention time, on the survival of SARS-CoV-2 in the aquatic environment. This requires the execution of tests aimed at assessing the virus vitality, practically almost never performed, until now.

However, some preliminary considerations about the environmental behaviour of the virus can be advanced. For example, Hart and Halden (2020) have quantitatively estimated the presence of the virus in the WW of infected areas starting from epidemiological data and assuming an exponential decay rate, substantially influenced by temperature, although its actual role, as well as possible synergistic effects with other factors should be further explored. Anyhow, this study shows a singular and innovative approach, which allows to predict the entity of WW contamination on a large scale, in areas, where the epidemics occurs with varying intensity.

Another interesting aspect is that SARS-CoV-2 belongs to the coronavirus family exactly like SARS-CoV. After the SARS epidemic of 2003–2004, some tests were conducted for assessing the virus survival in hostile environments. Wang et al. (2005) investigated the inactivation time in urban and hospital WW under different temperature conditions. In both cases at low temperatures (4 °C) the virus maintained its vitality up to 14 d, while it was inactivated only after 2d at higher temperatures (20 °C). The coronaviruses reduced survival at temperatures higher than 20 °C was also demonstrated by Gundy et al. (2009), who observed 99% HCoV-229E inactivation (23 °C) after 2.36 days and 1.85 days in urban WW after primary and secondary treatment, respectively. For SARS-CoV-2, some preliminary results were published by Rimoldi et al. (2020) which revealed that virus present in WW and river water collected in Milan did not exhibit cytopathic effect. In this case, only a limited number of samples was examined in a very short period of pandemic disease, therefore further research is required.

**Table 2:** *Literature results on presence of SARS-CoV-2 in wastewater. a: total number of samples not available, b: expressed as C_q_. n.a.: not available. [two-column fitting table]*

| References | Location | Number of<br>points examined | Period of<br>examination | Samples |  |
| --- | --- | --- | --- | --- | --- |
| | | | | Positive<br>(total<br>number of<br>samples) | $C_t$ value |
| Or et al. (2020) | Israel | 17 | 10/03/2020 -<br>21/04/2020 | 10 (26) | 32.76 - 38.5 |
| F. Wu et al. (2020) | U.S.A.<br>(Massachusetts) | 2 | 08/01/2020 -<br>25/03/2020 | 10 (14) | 33.87 - 38.39 |
| Rimoldi et al. (2020) | Italy | 3 | 14/04/2020 -<br>22/04/2020 | 4 (12) | n.a. |
| Ahmed et al. (2020) | Australia | 3 | 20/03/2020 -<br>01/04/2020 | 2 (9) | 37.5 - 39 |
| Medema et al. (2020) | Netherlands | 7 | 05/02/2020 -<br>16/03/2020 | 14 (24) | n.a. |
| Randazzo et al.,<br>(2020a) | Spain | 3 | 12/02/2020 -<br>14/04/2020 | 13 (15) | 34.00 - 37.84 |
| Randazzo et al.<br>(2020b) | Spain | 6 | 12/03/2020 -<br>14/04/2020 | 37 (72) | 34.00 - 40.00 |
| Wurtzer et al. (2020) | France | 3 | 05/03/2020 -<br>23/04/2020 | 100 % of<br>samples<br>positive <sup>a</sup> | n.a. |
| La Rosa et al.<br>(2020b) | Italy | 4 | 03/02/2020 -<br>02/04/2020 | 6 (12) | n.a. |
| Kocamemi et al.<br>(2020a) | Turkey | 9 | 21/04/2020 -<br>25/04/2020 | 7 (9) | 34.67 - 39.54<br><sup>b</sup> |

#### 4.1.2. How to remove it?

Although the coronaviruses survival in the environment can differ according to the specific type (Casanova et al., 2009; Wigginton et al., 2015), these indications allow to formulate some preliminary considerations. Based on the above-mentioned findings (Section 4.1.1.), SARS-CoV-2 might be present in WW, but its survival is likely extremely low, especially when the temperature of the WW remains stably above 20 °C. To date, the real influence of other parameters, like pH, sunlight and disinfectant agents, on survival time is still uncertain. Nonetheless, presuming a behaviour similar to that of other coronaviruses, this pathogen should rapidly be inactivated (WHO, 2020c).

Wang et al. (2005) observed that SARS-CoV can be completely inactivated in 30 minutes in the presence of residual chlorine greater than 0.5 mg L^−1^ or concentrations of chlorine dioxide equal to 2.19 mg L^−1^, resulting in an increased sensitivity of SARS-CoV towards chlorine in comparison to other pathogens such as *E.coli* and f2 phage. These results are confirmed by some preliminary tests on SARS-CoV-2 conducted *in vitro* by Chin et al. (2020) using 1:99 diluted household bleach. They observed that the virus was completely undetectable after 5 minutes of contact time.

This could suggest that the use of chlorine in plants would yield the complete inactivation of SARS-CoV-2. Although to date there is no evidence of COVID-19 transmission through sewage, either raw or treated (Lodder and de Roda Husman, 2020; WHO, 2020c), the WHO has recommended a disinfection stage in order to prevent any virus release into the receiving aquatic bodies (WHO, 2020c). Randazzo et al. (2020b) published a preliminary study on SARS-CoV-2 presence in WW after secondary and tertiary treatment. 11% of samples after conventional activated sludge resulted positive to SARS-CoV-2 RNA and 100% got negative after tertiary treatments (disinfection with NaClO, in some cases coupled with UV). Although this research does not clarify the influence of disinfectant dosage and contact time on virus survival and presents some limits such as the low number of samples, it shows that tertiary treatments already present in the majority of WWTPs could be enough to remove SARS-CoV-2. Besides, membrane bioreactors have shown promising results in virus removal, although the role of membrane materials and properties, of suspended solids and cake layers, of electrostatic interactions has to be further investigated (Gentile et al., 2018; Miura et al., 2015; Purnell et al., 2016).

### 4.2. SARS-CoV 2 and sewage sludge

#### 4.2.1. Is the presence of the virus in sewage sludge possible?

Given the presence of SARS-CoV-2 in the faeces and in the WW, the possible presence of the virus in the SS is arousing considerable interest (NEWEA, 2020; Núñez-Delgado, 2020). In particular, the attention concerns the possibility that the virus can survive the treatment phases and, when the SS is spread in agriculture, these could be a vehicle of contagion for the population. However, the search for SARS-CoV-2 RNA alone is not sufficient to determine any risk situations due to the spreading of the SS on the soil. The discovery of the nucleotide sequences of the virus is not in itself indicative of the virulence of the virus itself.

Although to date there are no a detailed literature on the presence of the virus in the SS, some preliminary results are available. Recently, during CoViD-19 outbreak in Turkey, Kocamemi et al. (2020) analysed primary and secondary sludge of 2 and 7 WWTPs, respectively. They found SARS-CoV-2 RNA in both types of sludge with comparable C_q_ (from 33.52 to 35.86). Although the results presented are very interesting since represent the first confirmation of presence of SARS-CoV-2 in SS, two main aspects still to be clarified. Firstly, the presence of the virus is not necessarily a prove that could be infective creating a risk for people. Secondly, in this study samples were taken immediately after the extraction from settling tanks and therefore the effect of treatments in the sludge line on SARS-CoV-2 presence and virulence were not considered. Peccia et al. (2020) analysed more than 40 samples taken daily during CoViD-19 outbreak and peak in Connecticut (U.S.A.). The most interesting point is that they not only tested the presence of SARS-CoV-2 in SS but also highlighted that RNA concentration can be used as indicator for the prediction of CoViD-19 spread evolution. It should be noted that in this experimentation the samples were taken immediately after the thickening phase, therefore the results shown do not necessarily imply that SARS-CoV-2 may be present in the SS at the time of the spread in agriculture. However, there are several aspects that the literature is not yet able to clarify, namely: (i) the impact of conventional and advanced treatments of the WWTPs sludge lines on the presence of SARS-CoV-2, (ii) the possible presence in the SS spread in agriculture (to date not demonstrated), and (iii) the virulence of SARS-CoV-2 in these contexts.

Even considering the other HCoVs, the literature data are extremely reduced to date. Bibby et al. (2011) found the presence of HCoV-229E and HCoV-HKU1 in SS even after the anaerobic digestion phase (35–37 °C, hydraulic retention time equals to 15 d) and mechanical dehydration. This study highlights the possibility that HCoVs can survive digestive phases in the mesophilic field, but it does not determine the virulence capacity of the viruses present in the SS. The samples were taken immediately after the treatment phases and therefore other factors that could accelerate the disappearance of the virus in the hypothesis in which they are spread on the soil such as for example pH variations, solar irradiation, and heat were not taken into consideration. HCoV-HKU1 was again identified by Bibby and Peccia (2013) in anaerobically digested SS. They expressed concern due to the high load of virus in treated SS that could be transmitted by the aerosols emitted during the land application. However, also in this case, the infectivity of the viruses, the influence of the anaerobic digestion temperature and meteorological conditions on the resistance of the virus were not defined.

As also recognized by U.S. National Research Council and Italian National Institute of Health (INIH, 2020; USNRC, 2002), the presence of the viruses in the SS is not directly indicative of a potential hazard of the matrix as an effective transmission capacity of the pathogen is not proven. Moreover, SARS-CoV manages to resist in the external environment for a limited time if subjected to heat and solar radiation (Darnell et al., 2004) and there are currently no studies on its possible survival on soil.

#### 4.2.2. How to remove it?

Generally, there are three main aspects that can improve the removal of viruses: (i) high retention time, (ii) high values of temperature and (iii) high/low values of pH. Recent studies show that also SARS-CoV-2 is particularly sensitive to these three aspects. Chin et al. (2020) have demonstrated how the resistance of the virus from 22 °C to 70 °C dropped from 7 d to 1 min, highlighting its sensitivity of this virus towards high values of temperature. This result is very similar to the previous findings in case of SARS-CoV by Rabenau et al. (2005), who observed greater stability compared to HCoV-229E in the external environment, but a rapid inactivation under temperature conditions of 56–60 °C. On the other hand, it has been shown that even significant changes in pH (from 3 to 10) would not seem to determine the disappearance of SARS-CoV-2 (Chin et al., 2020).

Although the studies on SARS-CoV-2 must be considered preliminary, as they are carried out only in vitro, they provide interesting insights that can enable the selection of the most appropriate treatments, in order to ensure complete inactivation of SARS-CoV-2 in the SS. Taking into account its sensitivity for temperature values greater than 50 °C, conventional and advanced biological thermophilic processes (e.g. anaerobic digestion (Leite et al., 2017) and aerobic/anaerobic membrane bioreactors (Collivignarelli et al., 2017), respectively), and thermal drying treatments (Richard, 2019) might presumably ensure the complete virus inactivation. Based on these first results, the use of mesophilic treatments could also be enough to determine the viral disappearance if associated with longer retention times.

However, none of the aforementioned studies on the influence of the different parameters on the inactivation of SARS-CoV-2 and SARS-CoV focus specifically on SS but it is very likely that, beside temperature (responsible for protein inactivation and extracellular enzymes activity increase), also the presence of bacteria, protozoa, metazoa, and, more generally, organic matter affect the virus survival (Gundy et al., 2009; Pinon and Vialette, 2018; Rzeżutka and Cook, 2004). Therefore, an increase in retention time might be proficuous. Anyway, owing to the total absence of literature regarding the presence and the influence of temperature, pH and retention time on the inactivation of SARS-CoV-2 in SS, further studies are recommended.

Despite the absence of literature on this topic, health authorities have issued precautionary guidelines. For example, to minimize any risk situations of reuse in agriculture, the Italian National Institute of Health (INIH, 2020) proposes “good practices” which provide for a minimum time of SS retention before reuse in agriculture inversely proportional to the temperature (Eq.1):

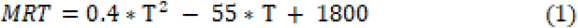

Where MRT is the minimum retention time (including treatment and storage times) in hours and T is the operating temperature (°C) (INIH, 2020). It should be noted that satisfying the MRT does not necessarily require significant changes in current sludge treatment and management practices. In fact, applying the formula in mesophilic conditions (30 – 35 ° C), it can be seen how the suggested precautionary MRT, before reuse in agriculture, is 15 – 21 days, therefore guaranteed by the retention time of an anaerobic digester for SS.

## 5. Tips and ideas for future research

From the analysis of the literature, the lack of data about the influence of parameters like temperature, pH and retention time on the survival of the virus in WW and SS is evident. Since *ad hoc* studies on SARS-CoV-2 are currently scarcely available, information can be inferred from experiments conducted in the past years on SARS-CoV, mostly *in vitro*. As confirmed by several authors (Carducci et al., 2020; Kitajima et al., 2020; La Rosa et al., 2020a), part of the lack of results on this issue is attributable to the difficulty in finding SARS-CoV-2 RNA within these particular matrices, as there is currently no officially recognized univocal method. Sewage and sludge contain hundreds of molecules (namely, fulvic and humic acids, fats, proteins, metal ions, detergents), liable to interfere with polymerase chain reaction: new techniques of molecular biology, such as the digital PCR and the next generation sequencing, might allow overcoming this issue. Furthermore, the adoption of different commercial kits for performing the PCR leads to the obtainment of results only partially comparable, owing to the variability of extraction efficiency (Mao et al., 2020; Sims and Kasprzyk-Hordern, 2020). Moreover, La Rosa et al. (2020b), regarding the monitoring of wastewater, highlight the inadequacy of the current techniques for concentrating and recovering non-enveloped enteric viruses. Therefore, identifying and sharing protocols from sample pre-treatment to viral particles quantification would allow simplifying, accelerating and stimulating the analysis of these matrices, before the issuing of a standardized technique.

The authors suggest evaluating the effect of the different parameters, possibly simultaneously, on the inactivation of SARS-CoV-2 in real matrices, like sewage, sludge, soil, freshwater (both surface and groundwater). A deeper knowledge of the actual occurrence and persistence of the infectious virus is mandatory. These aspects would also allow predict the behaviour of SARS-CoV-2 in the different phases of the WWTP, to identify possible exposure risks for workers. Furthermore, the possibility to track precisely the virus vitality (if any) from sewers to the WWTPs throughout the subsequent phases of treatment efficacy will help to optimize process conditions and to select more adequate technologies (with particular focus on disinfection).

Although WBE might reveal a precious tool for the early warning of future outbreaks and tracking the epidemic geographically and over time (Barcelo, 2020; Kitajima et al., 2020; Lodder and de Roda Husman, 2020; Sims and Kasprzyk-Hordern, 2020; F. Wu et al., 2020; Wurtzer et al., 2020) (Fig. 2) its thorough and diffused application still requires the acquisition of further information on crucial elements, which represent the input data of possible simulations. It is worth underlining the need of deeper studies about the virus shedding by infected asymptomatic and paucisymptomatic people, the actual stability of viruses and virions in sewage and, definitely, the possibility to exploit, in parallel, other biomarkers/pharmaceuticals to track the outbreak, in order to integrate the available information and obtain a more precise assessment for surveillance purposes.

**Fig. 2:**
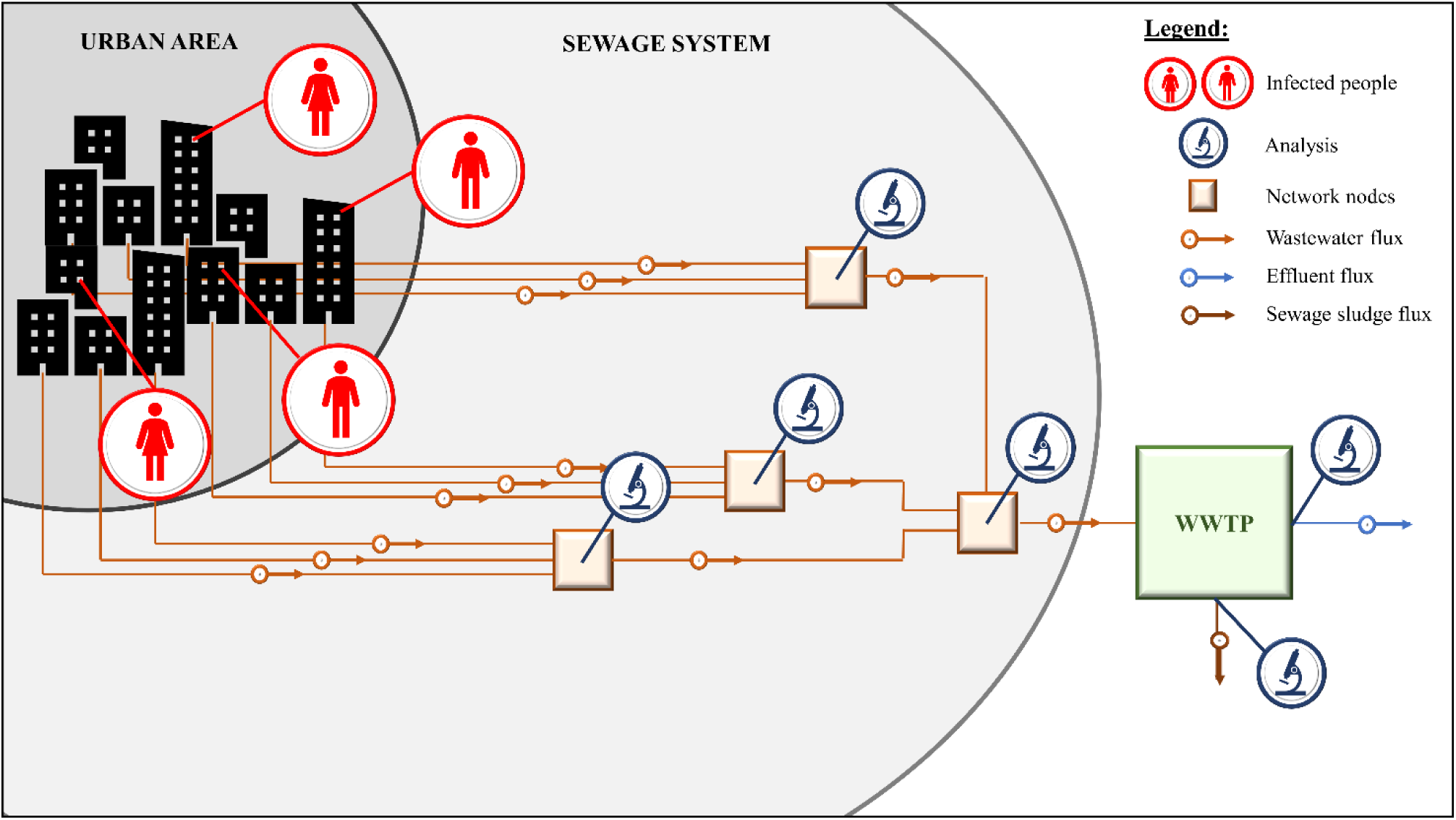
*Proposed scheme for monitoring the infection status of urban areas. [two-column fitting image]*

## Data Availability

Nothing to declare.

## Declaration of competing interest

The authors declare no conflicts of interest

## CRediT author statement

## Maria Cristina Collivignarelli

Methodology, Writing, Supervision. **Carlo Collivignarelli:** Methodology, Writing. **Marco Carnevale Miino:** Methodology, Writing, Visualization. **Alessandro Abbà:** Methodology, Writing, Visualization. **Roberta Pedrazzani:** Methodology, Writing, Validation. **Giorgio Bertanza:** Methodology, Writing, Supervision.

## Notes

### Competing Interest Statement

The authors have declared no competing interest.

### Funding Statement

No fundings received.

### Author Declarations

Nothing to declare.

